# Enhancing Access for Essential surgical and anesthesia care at Primary Healthcare Settings in Low-Resource Settings: A Scoping Review Protocol

**DOI:** 10.1101/2025.07.22.25331980

**Authors:** Siraj Ahmed Ali, Tadese Belayneh Melkie, Zewditu Abdisa Denu, Solomon Mekonnen

**Affiliations:** Dilla University College of Medicine and Health Sciences, Department of Anesthesiology, Dilla, Ethiopia; University of Gondar, College of Medicine and Health Science, School of Medicine, Department of Anesthesia, Gondar, Ethiopia; University of Gondar, College of Medicine and Health Science, Institute of Public Health, Gondar, Ethiopia

**Keywords:** surgical care, Anesthesia care, low-income countries, resource limited settings, access, barriers, enablers

## Abstract

**Introduction:** Access for safe, timely, and affordable surgical and anesthesia care is one of the integral components of universal health coverage with a significant global burden of treatable surgical diseases. Five billion people lack safe and timely surgical and anesthesia care globally, with more than 80% from low-income countries. The lancet commission on global surgery 9 in 10 people in low-income countries do not have access to essential surgical care. the problem associated with various factors, including infrastructure and manpower limitations. Recognizing the problem, there should be a tremendous strategy to give emphasis on improving access to safe, affordable, and timely surgical and anesthesia care. This scoping review aims to map existing evidence on perioperative anesthesia and surgical care access at the PHC level in LMICs to identify the barriers and enablers and suggest system-strengthening strategies for improving surgical care access in low-income countries.

**Methods:** The review will follow the Joanna Briggs Institute (JBI) methodology for scoping reviews and adhere to the PRISMA-ScR checklist. The review protocol has been registered at Open Science Framework (OSF). A comprehensive search strategy will be developed using a combination of key terms from various databases to generate published articles published between 2015 and 2025 and gray literature. Article selection and screening will use predefined and iteratively refined selection criteria based on the ‘Population–Concept– Context’ framework to independently screen titles and abstracts of citations from the search. The screening will be conducted by two independent reviewers with a predefined extraction tool. Disagreements will be solved by the two reviewers. The review will be reported in accordance with the PRISMA-ScR (Preferred Reporting Items for Systematic Reviews and Meta-Analyses Extension for Scoping Reviews) checklist.

## Introduction

Access for safe, timely and affordable surgical and anesthesia care is one of the integral components of universal health coverage (1). The global burden of treatable surgical conditions despite greater demand is significant. Despite of global recommendations and efforts, five billion lack access to safe, affordable surgical and anesthesia care with more than 80% from low income countries (2,3). This problem signifies that, by this limited access for essential surgical and anesthesia care, more than18 million deaths from preventable surgical conditions happening every year (4). The lancet commission on global surgery also found that 9 in 10 peoples in low income countries have not access for essential surgical care (5).

The problem of limited access for affordable, safe and timely surgical and anesthesia care in resource limited settings like sub-Saharan countries is dictated by various factors. There is significant infrastructure and critical human resource limitations. Less than half of the health facilities in those countries have not a capacity to provide essential and emergency services like cesarean section (6,7).

Recognizing the existing problem of inadequate surgical and anesthesia care access globally with significant proportion in the low and low middle income countries, there should be tremendous effort to give emphasis on improving access for safe, affordable and timely surgical and anesthesia care. There should also be evidence-based recommendations for determinants of the problem and workable strategies. The Lancet Commission on Global Surgery in recognition to the criticality of the issue targeted to identify barriers and opportunities for improvement in surgical care (2). The identification of barriers and opportunities for improving surgical and anesthesia care access especially in resource limited settings should be detected with feasible mapping of available evidences.

This scoping review aims to identify, explore, and map existing evidence on perioperative anesthesia and surgical care access at the PHC level in LMICs. It is anticipated that the results of this scoping review will inform government and policy makers on surgical services required to identify the barriers and enablers and suggest system strengthening strategies for improving surgical care access in low-income countries and indicates gaps for future research. A comprehensive exploration of the existing evidences for barriers, enablers, strategies, and policy frameworks is necessary to inform scalable interventions and guides future research works. Scoping reviews are appropriate for mapping broad, complex, and heterogeneous evidence areas.

## Objective

This scoping review aims to map existing evidence on access to essential surgical and anesthesia care, identify enablers and barriers and strategies to integrate surgical care into primary healthcare settings of low-income countries.

### Review Questions

- What is the current state of surgical and anesthesia service access at PHC levels in LMICs?
- What are surgical and anesthesia services available in PHC facilities in LMICs?
- What are key barriers and enablers for the delivery of essential surgical and anesthesia care at PHC facilities of LMICs?
- What strategies, innovations, or models of care have been implemented to improve access and the community engagement in improving access?
- What gaps exist in policy, systems, and research that are relevant to Ethiopia?

## Methods

This protocol is for a systematic scoping review of literature reporting on essential surgical and anesthesia care available in primary health care facilities, barriers and enablers, in LIMCs/sub-Saharan Africa. A scoping review method is selected as it aims to outline different types of evidence on the area of interest and the gaps for further research. The proposed review will be guided by the methodological framework by Arksey and O’Malley’s methodological framework, with amendments to this framework by Levac et al (8) or Joanna Briggs Institute (JBI). The Arksey and O’Malley framework is made generally up of six stages: (I) identifying the research question; (II) identifying relevant studies; (III) study selection; (IV) charting the data; (V) collating, summarizing and reporting results; and (VI) consultation. In this review we will follow five steps (I) identifying the research question, (II) identifying relevant studies, (III) selection of eligible studies, (IV) charting the data, and (V) collating and summarizing and reporting the results. Quality appraisal will not be done as this review aims to map all research activities in this field. The review protocol has registered at Open Science Framework (OSF) at the link https://archive.org/details/osf-registrations-qyrvd-v1 with the registration DOI: https://doi.org/10.17605/OSF.IO/QYRVD.

### Step I: Identifying the research question

Timely and affordable access for essential surgical and anesthesia care is an integral part of universal health coverage(9). We want here will be to explore how access is determined in different settings of low income and low middle income countries. our objective is to determine what is known in the literature about essential surgical and anesthesia care in low and low middle income countries and Sub-Saharan Africa. The review will focus on the following questions;

→ What is the current state of surgical and anesthesia service access at PHC levels in LMICs?
→ What are surgical and anesthesia services available in PHC facilities in LMICs?
→ What are key barriers and enablers for the delivery of essential surgical and anesthesia care at PHC facilities of LMICs?
→ What strategies, innovations, or models of care have been implemented to improve access and the community engagement in improving access?
→ What gaps exist in policy, systems, and research that are relevant to Ethiopia?

This study will use the PCC format to align the study selection with the research question. Please describe here (the below population, concept and context).

#### Box 1

**Inclusion criteria**

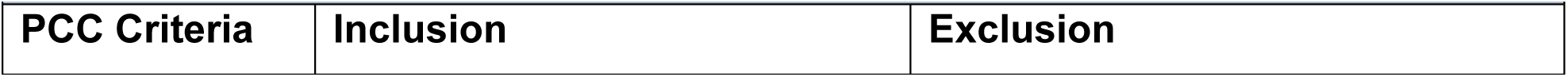
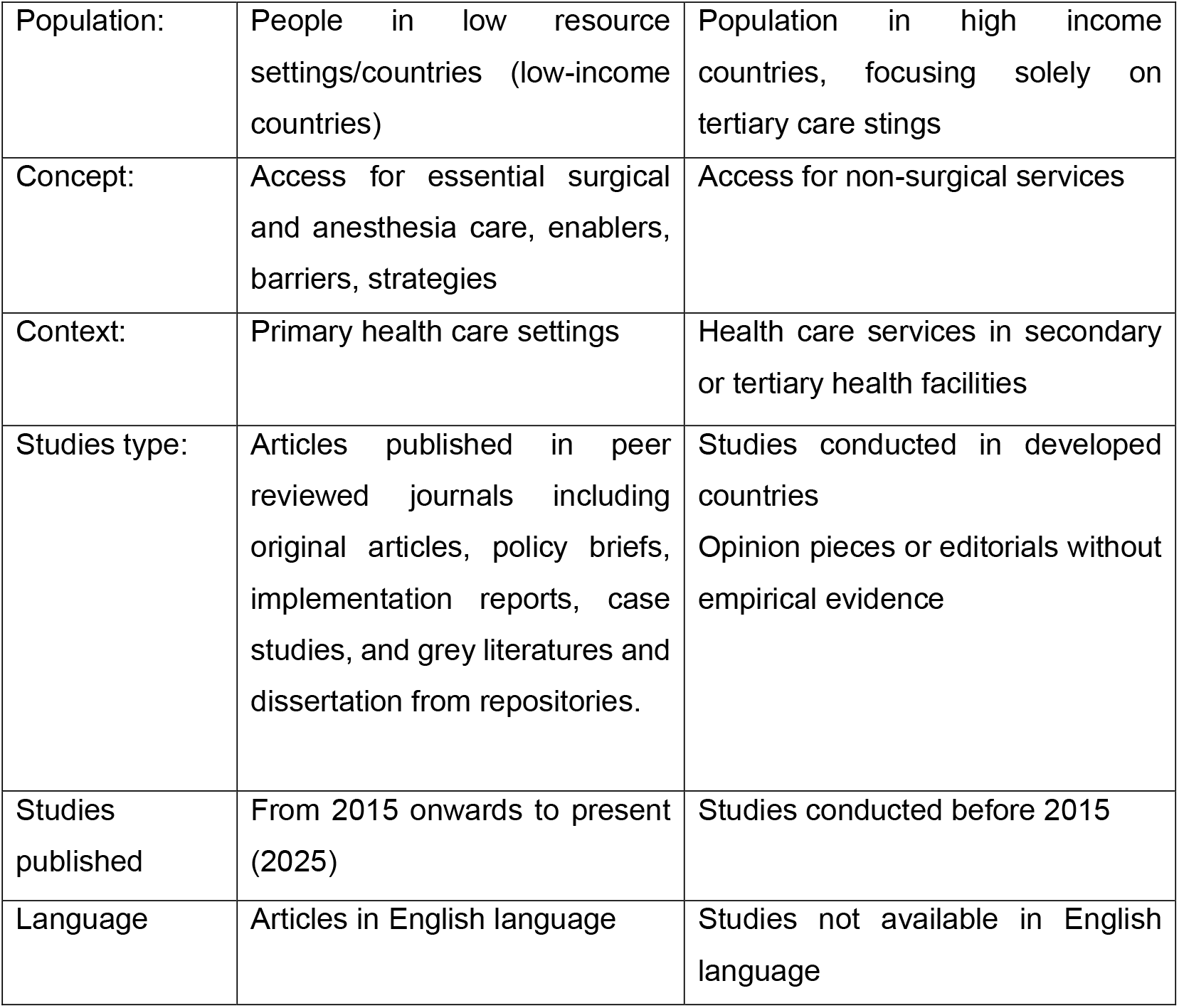

| Box 1: Inclusion criteria |  |  |
| --- | --- | --- |
| PCC Criteria | Inclusion | Exclusion |
| Population: | People in low resource settings/countries (low-income countries) | Population in high income countries, focusing solely on tertiary care settings |
| Concept: | Access for essential surgical and anesthesia care, enablers, barriers, strategies | Access for non-surgical services |
| Context: | Primary health care settings | Health care services in secondary or tertiary health facilities |
| Studies type: | Articles published in peer reviewed journals including original articles, policy briefs, implementation reports, case studies, and grey literatures and dissertation from repositories. | Studies conducted in developed countries<br>Opinion pieces or editorials without empirical evidence |
| Studies published | From 2015 onwards to present (2025) | Studies conducted before 2015 |
| Language | Articles in English language | Studies not available in English language |

### Step II: Identifying relevant studies

A search will be conducted for published and unpublished (grey) literature as per the developed search inclusion criteria based on the PRISMA recommendation of Population-Concept-Context framework (Box 1: selection criteria) on the research area in the following electronic databases: PubMed, Scopus, Web of Science, CINHAL, Global Index Medicus. Studies published since 2015 will be identified in the above data bases with the keywords or Medical Subject Headings (MeSH) terms *“surgical care”, “anesthesia care”, “primary health care”, “low middle income countries”, “low-income countries”*, “limited resource setting” with Boolean operators (“surgical” OR “perioperative” OR “anesthesia”) AND (“access” OR “barriers” OR “utilization”) AND (“low-income countries” OR “low-middle-income countries” OR “Limited resource setting”). To maximize the number of articles retrieved for review in the area, potentially relevant grey literature will be identified through targeted searches of dissertations/theses the ministry of health websites, university dissertation data bases to include important literatures.

The search strategy will be piloted to check the appropriateness of keywords and databases at early stage of literature search. A hand search will be also conducted of the references of the included studies to identify potentially relevant literature.

#### Box 2

**Search terms**

Surgical care

Anesthesia care

Perioperative care

Access

Low-income countries

Low middle income countries

Barriers

Enablers

Strategies

### Step III: Selection of eligible studies

Studies included in the scoping review will be based on the inclusion criteria developed by the reviewers (SA and TB), Box 1: showed selection criteria. The literatures Title and abstract screening will be guided by the PCC framework. Study selection will proceed in the following steps for academic databases; starting with selection based on title and abstract screening by two independent researchers (SA and TB). Disagreements in this step will be resolved by the two researchers themselves. The next step will be reading the selected and agreed citations in full by the two researchers independently, disagreements here at this point will be resolved by the researchers in consultation with third and fourth researchers (ZA and/or SM). Exclusion will be explained for texts not included after full-text perusal in this second step. In the third step, recommended by JBI, the reference lists of the included citations will be hand searched for relevant publications. We will use Zotero to store and keep track of our citations at the different stages and also allow us to identify and remove duplicates. The extraction of articles will be managed with the PRISMA 2020 flow diagram for systematic reviews (10) and included studies for final review will be identified.

**Figure 1.**
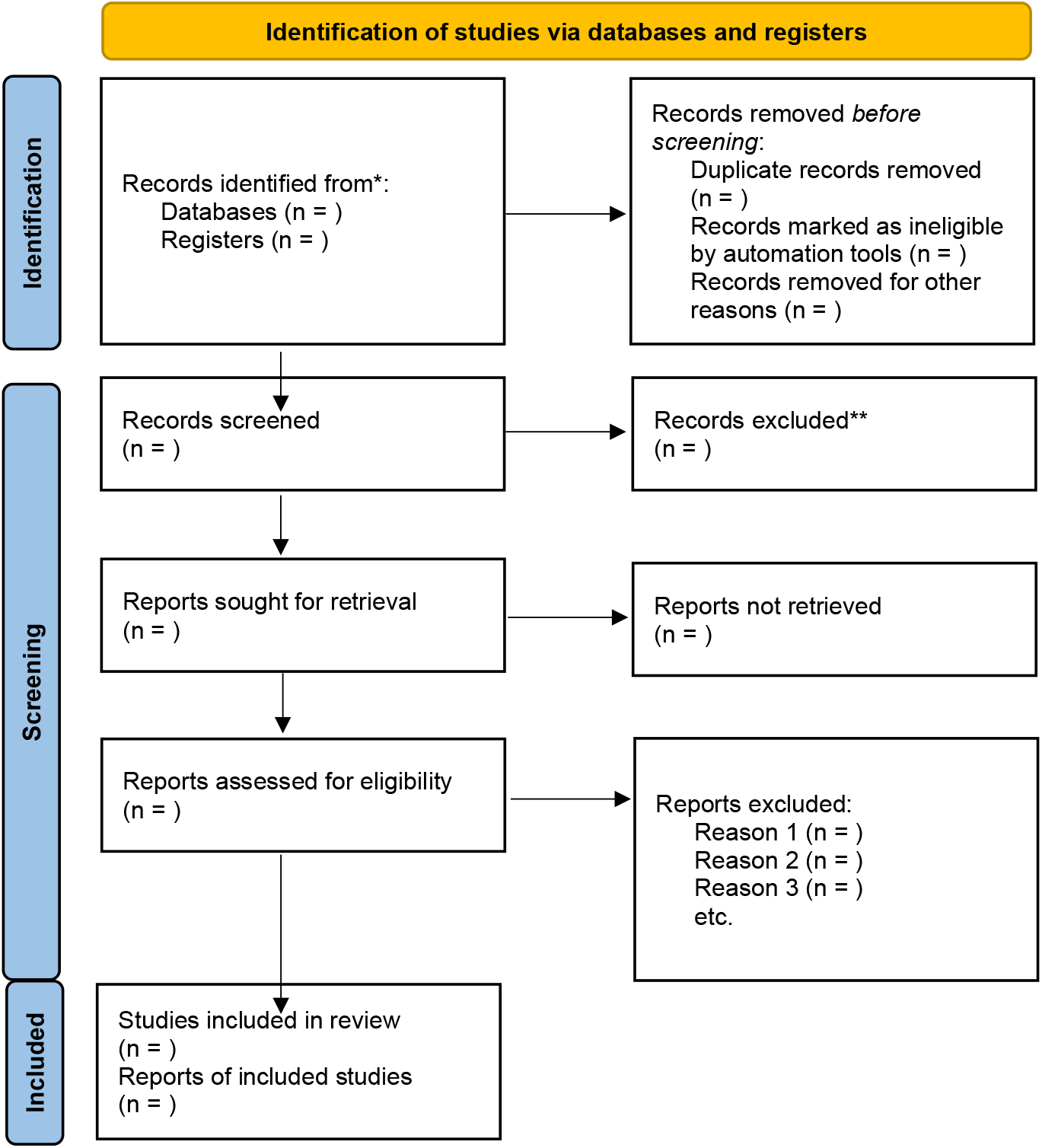
PRISAM 2020 flow diagram for systematic reviews.

### Step IV: Charting the data

Articles extracted from all databases including grey literatures will be summarized with data extraction form developed by researchers prior to conducting of literature search (Annex 1. Data extraction form). Authors will search data bases and various websites of relevant institutions based on the extraction form. Additional categories emerged during searching and charting will be added in the extraction form with additional columns when necessary. Two independents’ researchers (SA and TB) will work together to accomplish data extraction. Checkups and clearing will be conducting by other two researchers (ZA and SM).

A standardized data extraction form will be used to chart the following:

- Author(s), year, country
- Study design and setting
- Health facility level
- Focus area (workforce, policy, financing, service delivery)
- Key findings: barriers, enablers, models
- Reported evidence gaps
- Relevance to Ethiopian context

### Step V: Collating, summarizing, and report results

All eligible articles will be exported to the Zotero 7.0.21, duplication will be identified and removed. Titles and abstracts screening of all eligible articles will be conducted to determine the inclusion in to the review. The charted data will be analyzed thematically using tables. The table will show the results of the review used as bases for our discussion. Attempts will be made to obtain full texts of selected articles by title and abstracts using searching web with all authors. The two authors will conduct screening of the selected full text articles and the third reviewer will conduct further screening if there are significant discrepancies that cannot be resolved by discussion and consensus. The selection process will follow the recommendations of Preferred Reporting Items for Systematic Reviews and Meta-Analyses Extension for Scoping Reviews (PRISMA-ScR) checklist (Annex 2) (11) and be mapped using the PRISMA-P chart.

Extracted data will be synthesized thematically and presented in tables, charts, and narrative summaries. Themes may be structured using health systems frameworks (e.g., WHO building blocks). Gaps will be clearly highlighted based on the findings we get.

## Data Availability

All data produced in this review will be available upon the reasonable request.

## ETHICS AND DISSEMINATION

The results of this scoping review will be disseminated through a peer-reviewed publication and various conferences for targeting stakeholders to whom surgical anesthesia care access enhancement is relevant. The results will also be used to inform further research, determining various factors for surgical and anesthesia care access including barriers and enablers resource limited countries. Since this is a review of existing literature, no ethical approval is required. However, the protocol can be registered with open sciences framework (OSF).

## Contributors

SA, TB, ZA and SM were involved in the manuscript development in the following roles; conception and design of work, analysis of data for work, and drafting or revising critically for important stage of academic work, and final approval of the version to be published; and agreement to be accountable for all aspects of the work in ensuring that questions related to the accuracy or integrity of any part of the work are appropriately investigated and resolved.

## Acknowledgments

This review protocol has prepared to conduct scoping review for the course *Scoping Review of literatures*, part of the PhD study course work. Especial gratitude for university of Gondar college of Medicine and Health sciences for the opportunity to pursue this doctoral program and Dilla University for its provision of academic scholarship. Special thanks are extended to my primary supervisor, Dr. Tadese Belayneh, and co-supervisors, Dr. Zewditu Abdisa, and Prof. Solomon Mekonnen for their valuable feedback and constructive support.

## Patient Consent for publication

Not required.

## Annexes Annex 1: Data extraction form template

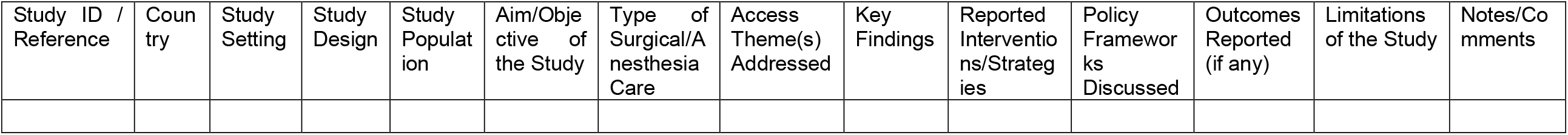

## Annex 2 Preferred Reporting Items for Systematic reviews and Meta-Analyses extension for Scoping Reviews (PRISMA-ScR) Checklist template

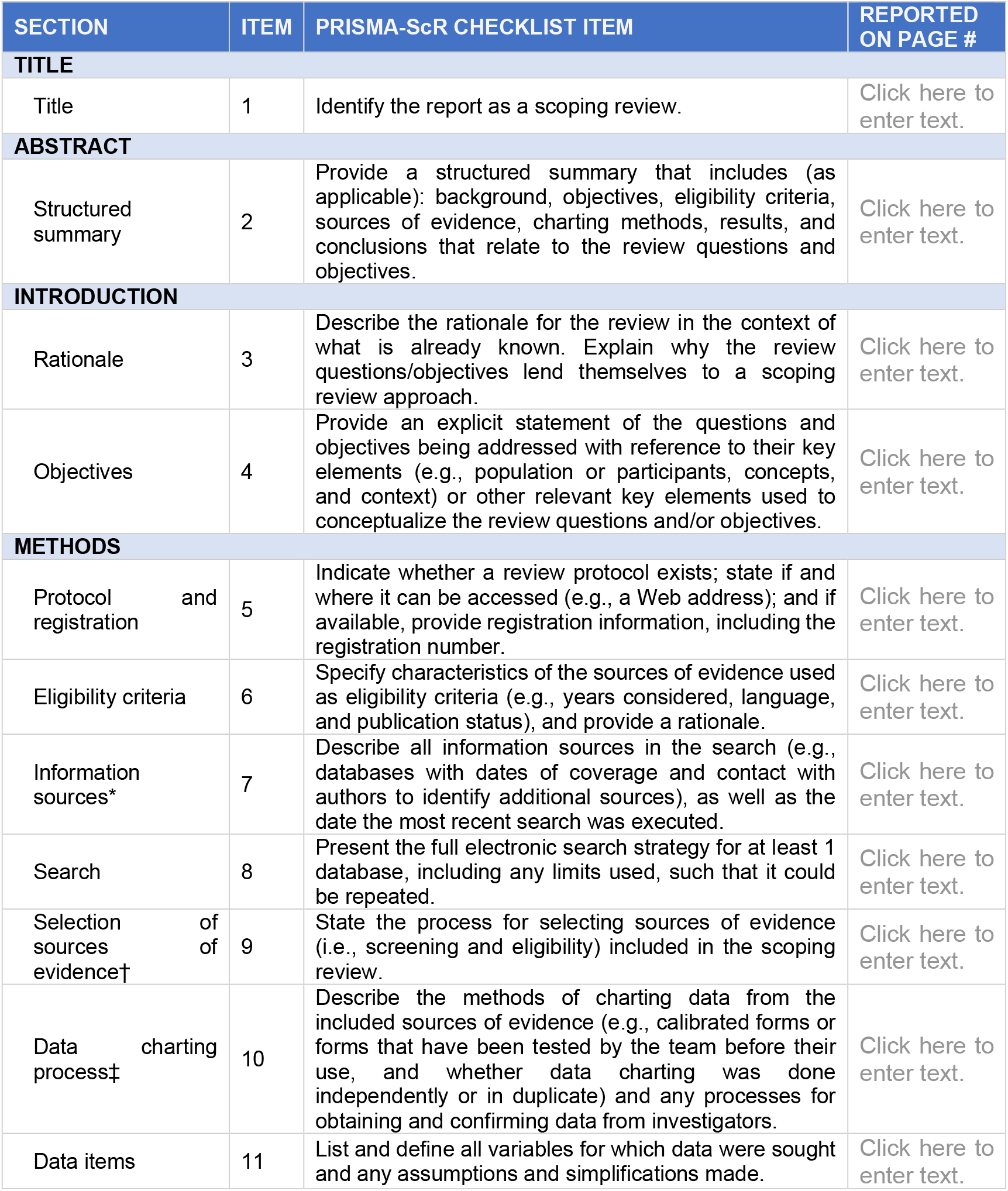
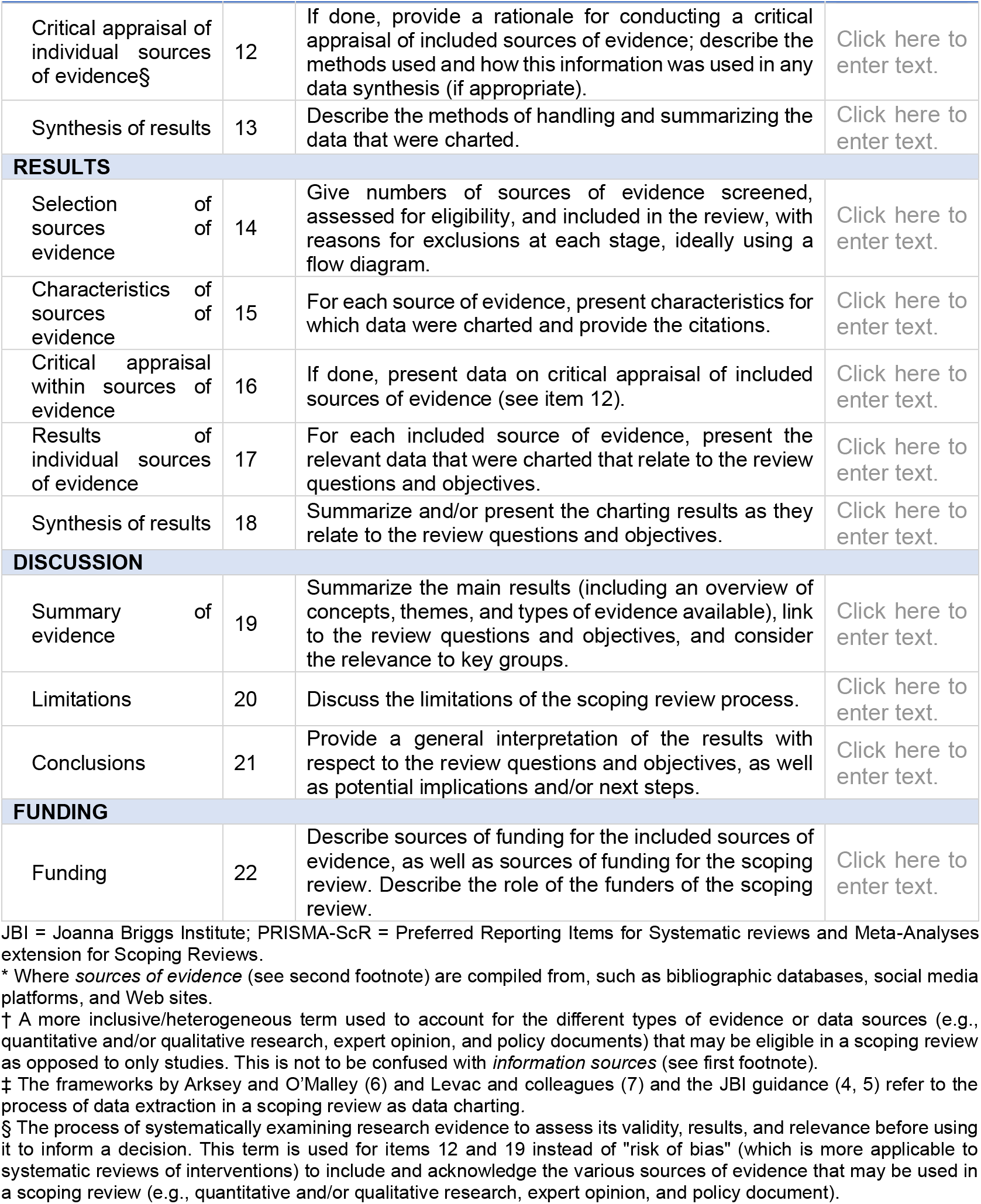

*From:* Tricco AC, Lillie E, Zarin W, O’Brien KK, Colquhoun H, Levac D, et al. PRISMA Extension for Scoping Reviews (PRISMAScR): Checklist and Explanation. Ann Intern Med. 2018;169:467-473. doi: 10.7326/M18-0850

